# Aerobic capacity predicts skeletal but not cardiac muscle damage after a full distance Ironman triathlon - the Iron(wo)man-study

**DOI:** 10.1101/19001149

**Authors:** Tom Danielsson, Jörg Carlsson, Lasse Ten Siethoff, Jonas Ahnesjö, Patrick Bergman

## Abstract

**Purpose:** This study examines the association between aerobic capacity and biomarkers of skeletal- and cardiac muscle damage among amateur triathletes after a full distance Ironman.

**Methods:** Men and women (N=55) were recruited from local sport clubs. One month before an Ironman triathlon, they conducted a 20m shuttle run test to determine aerobic capacity. Blood samples were taken immediately after finishing the triathlon, and analyzed for biomarkers of cardiac- and skeletal muscle damage. Regression models examining the association between aerobic capacity expressed in both relative terms (mLO2*kg-1*min-1) and absolute terms (LO2*min-1) controlled for weight and were fitted.

**Result:** A total of 39 subjects (26% females) had complete data and were included in the analysis. No association between aerobic capacity and cardiac muscle damage but a significant negative association between aerobic capacity and skeletal muscle damage was observed. This association was independent of how aerobic capacity was expressed, although the model with aerobic capacity expressed in absolute terms and controlled for weight resulted in slightly higher r^2^ values, than when aerobic capacity was expressed in relative terms.

**Conclusion:** A negative association between aerobic capacity and skeletal muscle damage was seen but despite the well-known cardio-protective health effect of high aerobic fitness no such association could be observed in this study.

## Introduction

Ultra-endurance races such as the Ironman triathlon (3.8 km swimming, 180 km cycling and 42.2 km running) has been on the rise since the mid-1980s and the proportion of women participating is currently around 15 % [1]. Even though physical activity in general has positive health effects, ultra-endurance races, such as an Ironman triathlon, lead to a rise in biomarkers of skeletal- and cardiac muscle damage [2]. However, most of the previous studies in the area have been conducted on elite athletes and the physiological responses to such performances may not be applicable to less trained, amateur athletes. For example, we have previously shown that both female and male amateur athletes have elevated cardiac troponin T (cTnT) well above the clinical reference limit for myocardial infarction after a Ironman triathlon [2] and that another marker for myocardial damage, cardiac specific myosin heavy chain *α* (MHC-*α*) follows a two peaked increase post-race [3]. However, the magnitude of the changes were greater in our sample compared to other studies on elite athletes [4]. Moreover, we were unable to explain the variation of the skeletal and cardiac muscle damage with our models, which included sex, age, body composition and finishing time (r2 values ranged from 0-22 %).

In order to better understand the health implications of the damage observed after ultra-endurance races, and to identify strategies for prevention, it is important to find variables that improve the model of skeletal- and cardiac muscle damage prediction. In this regard, maximum aerobic capacity is an interesting candidate since it has been well documented that having a high aerobic capacity is protective against cardiovascular disease in the general population [5]. In addition, the physiological factors that leads to high aerobic capacity could also protect the athlete from cardiac and skeletal muscle damage. For instance, higher aerobic capacity has been linked to left ventricular [6] and cardiac myocyte 6 adaptations that might minimise cardiac muscle damage. Higher aerobic capacity should in theory also limit skeletal muscle damage since it is correlated to training volume [7] and intensity [8], which lead to structural adaptations that increase the skeletal muscles’ tolerance to stress [9].

There are several ways of expressing aerobic capacity and the most commonly used is oxygen uptake expressed per body weight, i.e. mLO2*kg^−1^*min^−1^ However, expressing aerobic capacity relative to body weight also implies a proportional relationship between the two variables; if there is no such relationship the ratio becomes invalid [10]. Expressing aerobic capacity as a ratio complicates any interpretation of which of the two variables, or if the ratio itself, correlates with the outcome. Entering the two variables that make up the ratio, i.e. oxygen consumption (LO2*min^−1^) and weight (kg), as separate variables when performing statistical analyses is suggested as a solution to overcome the ratio problem [11]. In this study we examined the association between aerobic capacity and skeletal- and cardiac muscle damage after a full distance Ironman in a sample of amateur triathletes. Aerobic capacity was expressed both relative to kg body weight (mLO2*kg^−1^*min^−1^), as well as in absolute terms (LO2*min^−1^) corrected for weight, in order to see if one is a better predictor than the other.

## Methods

### Study sample

In all, 55 subjects, who had signed up to participate in an Ironman triathlon in Kalmar, Sweden, were recruited from local sport clubs associated with the Swedish Triathlon Federation. Because our sample was not selected at random, there is a risk of selection bias and therefore, we tested for differences regarding age and finishing time from the overall population of Ironman participants (i.e. average obtained from 41,000 finishers). There were no significant differences in either age (p=0.590, one sample t-test) or finishing time (p=0.780, one sample t-test) between our sample and the overall Ironman-population.

### Ethical considerations

This study received ethical approval from the regional ethical committee in Linköping (Dnr 2016/86-31). Signed informed consent was obtained from all participants.

### Procedure

Approximately one month prior to the Ironman triathlon, all subjects were invited to conduct a 20m shuttle run test to estimate their aerobic capacity. In total, 42 subjects performed the test. On the same occasion, the subjects were measured for weight and percentage of body fat (%BF) using bioelectrical impedance analysis (BIA) on a Tanita BC-545 Body Composition Analyzer (Tanita, Inc., Tokyo, Japan). Height was measured on a SECA 217 stadiometer (SECA medical measurements and scales, Hamburg, Germany).

One week before and within 15 minutes after the race, blood samples were drawn from an antecubital vein by registered nurses. The samples were transferred to the local hospital to be analysed within two hours of being drawn, with the exception of MHC-*α* which was stored at -80^°^C and analysed within two months after the race was finished. As markers of skeletal muscle damage, the blood samples were analysed for creatine kinase (CK; ref. < 1.9 *μ*kat/L) and myoglobin (MG; ref. <72 *μ*g/L). Cardiac muscle damage was analysed using cardiac troponin T (cTnT; ref. < 14 ng/L) and the novel marker MHC-*α* (no available reference values) MHC-*α* is occurring predominantly in atrial muscle but as well in ventricular muscle [12]. N-terminal prohormone of brain natriuretic peptide (NT-proBNP; ref. < 300 ng/L) was analysed as a marker for cardiac volume overload. Levels of myoglobin cTnT and NT-proBNP were analyzed using an electrochemiluminiscience immunoassay on automated analyzers (Cobas e411, Roche Diagnostics GmbH, Mannheim, Germany). CK was analyzed using the multiple-point dry chemistry method (Vitros 5.1 FS Ortho Clinical Diagnostics, Ortho-Clinical Diagnostics, Johnson-Jonson Company, Rochester, USA). The laboratory methods used were standard hospital procedures and were analysed at a laboratory accredited according to Sweden’s national accreditation body, Swedac (www.swedac.se). MHC-*α* concentration was measured with a commercially available ELISA (Cloud-Clone, Houston, Texas, USA) with no reported cross reactivity with other myosins. The biomarkers were chosen because they are commonly used in clinical practice, they are based on recommendations for sports medicine [13] and they allow for comparison with previous research in the field.

### Statistics

All analyses were performed using IBM SPSS version 24. Descriptive data are presented as mean ± standard deviation (SD) or as median an 25^th^-75^th^ percentile for highly skewed variables. We performed a drop-out analysis in which we investigated if there were any differences in baseline characteristics between those with and without data on aerobic capacity. In the drop-out analysis, independent samples t-test were conducted. In order to examine the potential influence on aerobic capacity on skeletal- and cardiac muscle damage, we created two sets of models, one in which aerobic capacity is expressed relative to weight (mLO2*kg^−1^*min^−1^) and one in which aerobic capacity is expressed in absolute terms (LO2*min^−1^). Both sets were analysed in both crude and adjusted analyses. The adjusted models were adjusted for baseline values of the biomarkers, sex, %BF and age of the participants. When aerobic capacity, in absolute terms, was analysed we also entered weight in the model. Furthermore, in the adjusted analyses, a stepwise regression with two steps was fitted. This was done to investigate the independent contribution of aerobic capacity on the dependent variables, i.e. the change in the adjusted values of coefficient of determination (r2). Despite several of the variables being correlated with each other, no sign of severe multicollinearity was observed in any of the models (all variance inflation factors < 5). If the assumption of heteroscedasticity of the residuals was not satisfied, the dependent variables were log transformed.

## Results

In total, 39 finished the race and provided baseline data. On average they had trained for 11.2 ±3 hours/week and had finished a median (25th-75th percentile) 8 (3-19) ultra-endurance races prior to this one. The descriptive data and baseline biomarker levels stratified for sex is presented in table 1. There were no differences between those with or without baseline assessment data in any of the variables shown in table 1 (all p>0.05).

**Table 1:** Descriptive statistics and baseline biomarker levels for men and women and the total sample among those who finished the race and had complete data. P-values are from an independent samples t-test for sex differences

|  | Male (n=29) | Women (n=10) | Total (n=39) |  |
| --- | --- | --- | --- | --- |
| | Mean ± SD | Mean ± SD | Mean ± SD | p |
| mLO2*kg<sup>-1</sup>*min<sup>-1</sup> | 51.5 ± 6.1 | 45.5 ± 5.2 | 49.9 ± 6.4 | 0.009 |
| LO2*min<sup>-1</sup> | 4.0 ± 0.6 | 2.8 ± 0.5 | 3.7 ± 0.8 | <0.001 |
| Age | 42.7 ± 10.6 | 37.8 ± 9.6 | 41.4 ± 10.4 | 0.208 |
| Weight | 77.8 ± 7.1 | 61.6 ± 5.4 | 73.7 ± 9.8 | <0.001 |
| Body fat (%) | 14.2 ± 3.9 | 22.5 ± 4.9 | 16.3 ± 5.5 | <0.001 |
| Cardiac Troponin T (ng/L) | 9.2 ± 4.8 | 6.7 ± 4.4 | 8.6 ± 4.8 | 0.159 |
| Myosin Heavy Chain-α (ng/L) | 1.1 ± 0.4 | 1.6 ± 0.7 | 1.3 ± 0.5 | 0.007 |
| NT-proBNP (ng/L) | 54.2 ± 17.4 | 59.2 ± 21.4 | 55.5 ± 18.4 | 0.466 |
| Creatine Kinase (μkat/L) | 3.8 ± 4.6 | 1.9 ± 0.9 | 3.3 ± 4.1 | 0.216 |
| Myoglobin (μg/L) | 40.9 ± 29.2 | 24.9 ± 5.7 | 36.8 ± 26.2 | 0.097 |

In the unadjusted analyses, a negative association between aerobic capacity (mLO2*kg^−1^*min^−1^) and both biomarkers for skeletal muscle damage and MHC-*α* was observed. No association between aerobic capacity in absolute terms (LO2*min^−1^) and the marker for skeletal muscle damage while a negative association for MHC-*α* were found (Table 2).

**Table 2:** Bivariate regression coefficients for aerobic capacity expressed both in relative (mLO2*kg^−1^*min^−1^)) and absolute terms (LO2*min^−1^)

|  | Aerobic capacity<br>(mLO2*kg<sup>-1</sup>*min<sup>-1</sup>) |  |  | Aerobic capacity<br>(LO2*min<sup>-1</sup>) |  |  |
| --- | --- | --- | --- | --- | --- | --- |
|  | Beta | 95% CI | r<sup>2</sup> | Beta | 95% CI | r<sup>2</sup> |
| Cardiac Troponin T (ng/L) | 0.01 | -0.02 - 0.05 | -0.012 | 0.04 | -0.28 - 0.36 | -0.025 |
| Myosin Heavy Chain-α (ng/L) | -0.05 | -0.08 - -0.01 | 0.104 | -0.60 | -0.89 - -0.31 | 0.300 |
| NT-proBNP (ng/L) | -0.03 | -0.06 - 0.01 | 0.036 | -0.28 | -0.57 - 0.01 | 0.072 |
| Creatine Kinase (μkat/L) | -0.06 | -0.10 - -0.02 | 0.203 | -0.32 | -0.65 - 0.02 | 0.068 |
| Myoglobin (μg/L) | -158.6 | -283.3 - -33.9 | 0.129 | -600.1 | -1722.5 - 522.4 | 0.005 |

In the unadjusted analyses, a negative association between aerobic capacity (mLO2*kg^−1^*min^−1^) and both biomarkers for skeletal muscle damage and MHC-*α* was observed. No association between aerobic capacity in absolute terms (LO2*min^−1^) and the marker for skeletal muscle damage while a negative association for MHC-*α* were found (Table 2).

In the adjusted analysis, a stepwise regression with two blocks was used to estimate the unique effect of aerobic capacity on the biomarkers (table 3). Aerobic capacity relative to weight (mLO2*kg^−1^*min^−1^) contributed significantly to the model predicting skeletal muscle damage. For myoglobin, adding aerobic capacity (mLO2*kg^−1^*min^−1^) increased the adjusted r2 from 0.026 to 0.210 (F: 8.927, p=0.005) and for CK the adjusted r2 increased from -0.015 to 0.267 (F: 13.778, p=0.001).

**Table 3:**
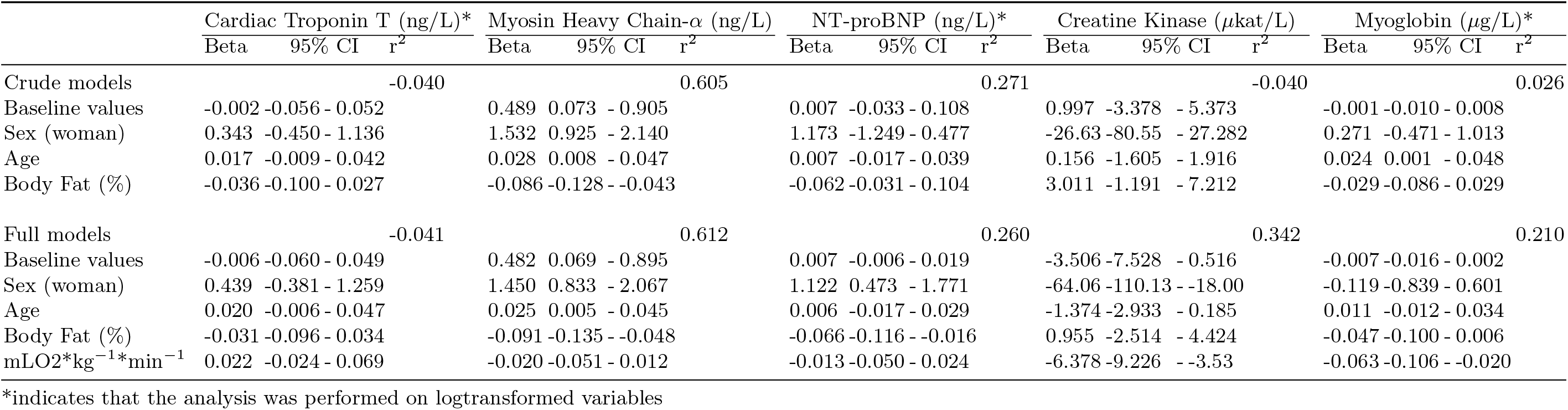
Regression models for the association between aerobic capacity expressed relative to body weight (mLO2*kg^−1^*min^−1^)) and the biomarkers

A similar picture regarding the independent association between aerobic capacity and the biomarkers was seen when aerobic capacity was entered in absolute terms (LO2*min^−1^) in the models (table 4). For myoglobin, the explained variance increased significantly from an adjusted r2 value of 0.07 to 0.227 (F: 10.386, p=0.003) and for CK from an adjusted 2 value of -0.029 to 0.281 (F: 15.215, p<0.001) when adding aerobic capacity to the models.

**Table 4:**
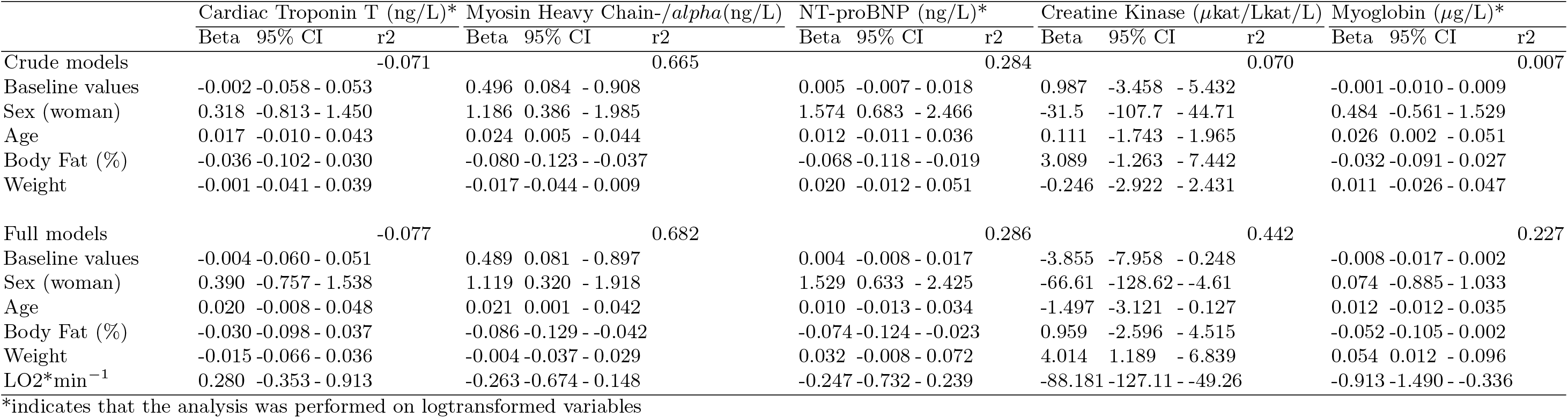
Regression models for the association between aerobic capacity expressed in absolute terms (LO2**min^−1^)) and the biomarkers

## Discussion

The aim of this study was to investigate the association between aerobic capacity and biomarkers of skeletal and cardiac muscle damage after a full distance Ironman in amateur athletes. We observed that increased aerobic capacity was associated with decreased skeletal muscle damage, while no evidence was found for an association with cardiac muscle damage. These relationships were independent of how aerobic capacity was expressed.

### Cardiac muscle damage

The absence of an association between aerobic capacity and markers for cardiac muscle damage was surprising given the well know cardio-protective health effects of having a high aerobic fitness. We have previously reported that nearly all of the athletes showed pathological values of cTnT (98%) and NT-proBNP (89%) after finishing the race [3] but the clinical value of these observations are under debate. The debate centers on whether the elevated biomarkers seen after ultra-endurance races reflect true pathological changes or if they are a part of the normal physiological response [14, 15]. Given that cTnT leaks into the bloodstream even after rather modest efforts such as after one hour of spinning [16] it might reflect myocyte stress rather than damage in a non-pathological population. For this reason, we included the novel marker MHC-*α*. While cTnT is a small molecule (37 kDa), this is much larger (224 kDa) and an increase after exercise could hardly be explained by passage through an intact cardiac cell membrane or as a normal physiological response. However, as a diagnostic marker, MHC-*α* has not been thoroughly studied and there are no, with modern technology, established reference values.

One possible explanation for the lack of association between aerobic capacity and cardiac muscle damage may be that subjects with high aerobic capacity exercise at higher aerobic power outputs. Higher aerobic power outputs demand higher cardiac outputs, which leads to increased cardiac stress. In fact, the rate of perceived exertion is a reliable predictor of training stress [17] which implicates that athletes automatically adapt the aerobic output to the aerobic capacity. The lack of positive association between aerobic capacity and cardiac damage could therefore be seen as indirect evidence that aerobic capacity actually is protective of cardiac damage if the required time and power output is constant. A better predictor for cardiac damage could be the relative intensity, i.e the working heartrate divided by the maximum heartrate which is a commonly used variable to describe intensity and load in endurance athletes. The importance of relative intensity as a predictor of cardiac damage is strengthen by the fact that higher intensity but for shorter duration leads to a greater cTnT release [18]. Measurement of the athletes working and maximal heartrates should therefore be included in future research. Another factor that should be considered for future research is the indirect measure of aerobic capacity. The 20m shuttle-run is provides reasonably good estimates of aerobic capacity [19] and is a useful field test since several subjects can be tested simultaneously at a low cost. However, it includes a lot of anaerobic accelerations and decelerations that might become a limiting factor for ultra-endurance adapted athletes. This may have affected the accuracy of the models in this study.

### Skeletal muscle damage

The association between aerobic capacity and skeletal muscle damage observed in this study could be due to several factors. Having a higher aerobic capacity is correlated to training volume [7] and intensity [8], which leads to structural adaptations that increase the skeletal muscles? tolerance to stress [9]. Another aspect that has been linked to skeletal muscle damage, is eccentric (breaking) muscle actions [20]. Eccentric muscle action occurs during running, but not during swimming or cycling. In this context, the inter-individual difference in running economy may be the most interesting factor in explaining why we observe a negative association between aerobic capacity and skeletal muscle damage post-race. Running economy is defined as the energy demand for a given velocity of submaximal running and is influenced by several factors, including ground reaction forces, muscle-tendon properties [21] and body weight [22]. Therefore, those with lower aerobic capacity may also have a poorer running economy, i.e., too much movement in the vertical direction. This results in higher ground reaction forces [23] with a subsequent increase in eccentric muscle actions and more knee-joint-movement that may lead to skeletal muscle damage. The potential associations between aerobic capacity, running economy and skeletal muscle damage are further supported by the associations between markers of skeletal muscle damage and body weight (table 4) that were seen in this study. Quantifying the ground reaction force during a race could be done by equipping the athletes with accelerometers which have been shown to be able to measure ground reaction forces [24].

### Expressing aerobic capacity

In this study, we used two different approaches to analyse the association between aerobic capacity and the biomarkers. This was done out of concern that entering a ratio, in our case aerobic capacity expressed relative to body weight, in the model could be problematic for two reasons. Firstly, the ratio itself may be biased and could cause spurious associations between the ratio and the health related outcomes to be observed 10,11. Conversely, a ratio may obscure the true relationship between itself and the health related outcomes. This has been proven by Kronmal, who showed that for another commonly used ratio, the body mass index 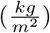, the ratio could appear unrelated to the health related outcomes, even though one or both of the variables making up the ratio was related to the same outcomes [25]. In our study, the two sets of adjusted regression models showed similar results, but they were not identical. In the models where aerobic capacity was expressed in absolute terms a negative association with the biomarkers for skeletal muscle was seen in combination with a positive association between weight and the biomarkers. In theory, this could lead to a situation in which the two variables would be cancelling each other out if they were combined to form a ratio. This was not the case in our study where aerobic capacity, independently of how it was expressed, was negatively associated with skeletal muscle damage. However, the adjusted r2-values were higher for the models with aerobic capacity expressed in absolute terms compared to relative terms, even if this difference was small indicating that a slightly clearer picture of the relationship between aerobic capacity and health related outcomes after a full distance Ironman-triathlon emerged.

### Strengths and limitations

In this study, the 20m shuttle-run was used to quantify the aerobic capacity of the triathletes. This is not a direct measure of aerobic capacity, which may have affected the accuracy of the models. However, it provides reasonably good estimates of aerobic capacity [19] and is a useful field test since several subjects can be tested simultaneously at a low cost. In contrast to most of the previous research in this field, we have included sample that is representative of the general participants and a relatively large proportion of women. This makes the results more relevant in a health context and increases the generalisability of our findings. The major limitation is that our study is an observational study, which limits our ability to draw conclusions regarding causal effects

### Practical application

The growing interest in ultra-endurance events warrants more research into the potential short and long-term health consequences of participating in such races. Most of the literature to date has focused on the acute changes in different biomarkers but information about underlying factors associated with those changes is scarce. In this study, we examined if aerobic capacity predicted the magnitude of cardiac- and skeletal muscle damage post-race. As we found no such association with cardiac muscle damage, our findings indicate that predictors of this remain to be identified. This is important information given the debate whether a marathon, triathlon or ultra-running repeatedly harms the heart by causing heart muscle cell necrosis. There is only circumstantial evidence on both sides of the debate of physiological versus pathological explanations of cTnT leakage. Neither side can so far conclusively prove what the mechanism of cTnT rise really is. On the one hand, it seems counterintuitive that just exercise should be the one and only scenario where increased plasma cTnT levels are not associated with increased risk of adverse outcomes. In all other situations even a subclinical rise of cTnT indicates a higher incidence of subsequent coronary heart disease, heart failure and death [26]. On the other hand, it seems equally unlikely that elite distance runners who maintain a high fitness over decades would have repeatedly sustained small myocardial infarctions [27]. Further studies using a longitudinal study design is needed to resolve this debate. To avoid excessive skeletal muscle damage, having a high aerobic capacity is beneficial, possibly linked to structural adaptations of the muscle-tendon unit associated with a higher training dose or with a better running economy.

### Conclusion

Aerobic capacity, independently of how it was expressed, was negatively associated with biomarkers for skeletal muscle damage after an Ironman triathlon, but not with biomarkers for cardiac muscle damage. To identify predictors of cardiac muscle damage, future studies could test other predictors, included more potential confounders or analyse more specific biomarkers, or include measures taken during the race if possible. The long term health effects of ultra-endurance races is an area or research which needs further attention.

## Data Availability

Data can be retrieved from the corresponding author at a reasonable request

## Acknowledgement

Thanks must go to the Kalmar Ironman organisers, the team of nurses who took the blood samples and of course the willing triathletes. We would also like to thank the laboratory team in Kalmar County Hospital and Marlene Norrby and Mohammad Ashikur Rahman from the Department of Chemistry and Biomedical sciences, Linnaeus University

## References

[1] R. Britt. How much time does it take to finish an ironman triathlon? (http://www.runtri.com/2011/06/how-long-does-it-take-to-finish-ironman.html?m=1), 2012.

[2] T. Danielsson, J. Carlsson, H. Schreyer, J. Ah-nesjö, L. Ten Siethoff, T. Ragnarsson, å. Tugetam, and P. Bergman. Blood biomarkers in male and female participants after an ironman-distance triathlon. PLoS One, 12(6):e0179324, 2017.

[3] T. Danielsson, H. Schreyer, H. Woksepp, T. Johansson, P. Bergman, A. Månsson, and J. Carlsson. Two-peaked increase of serum myosin heavy chain-alpha after triathlon suggests heart muscle cell death. BMJ Open Sport Exerc Med, 5(1):e000486, 2019.

[4] J. Carlsson, T. Ragnarsson, T. Danielsson, T. Johansson, H. Schreyer, A. Breyne, and P. Bergman. Hjärtmarkörer ökar efter intensiv motion - oklar klinisk betydelse - data från förstudie av kalmar ironwoman-studien visar på troponin t-värden som vid hjärtinfarkt. Lakartidningen, 113 [in swedish], 2016.

[5] S. N. Blair, 3rd Kohl, H. W., Jr. Paffenbarger, R. S., D. G. Clark, K. H. Cooper, and L. W. Gibbons. Physical fitness and all-cause mortality. a prospective study of healthy men and women. JAMA, 262(17):2395–401, 1989.

[6] J. Grewal, R. B. McCully, G. C. Kane, C. Lam, and P. A. Pellikka. Left ventricular function and exercise capacity. Jama-Journal of the American Medical Association, 301(3):286–294, 2009.

[7] S. A. Hawkins, T. J. Marcell, S. V. Jaque, and R. A. Wiswell. A longitudinal assessment of change in vo2max and maximal heart rate in master athletes. Medicine and Science in Sports and Exercise, 33(10):1744–1750, 2001.

[8] A. W. Midgley, L. R. McNaughton, and M. Wilkinson. Is there an optimal training intensity for enhancing the maximal oxygen uptake of distance runners? empirical research findings, current opinions, physiological rationale and practical recommendations. Sports Medicine, 36(2):117–132, 2006.

[9] P. M. Clarkson and M. J. Hubal. Exercise-induced muscle damage in humans. American Journal of Physical Medicine & Rehabilitation, 81(11):S52–S69, 2002.

[10] J. M. Tanner. Fallacy of per-weight and per-surface area standards, and their relation to spurious correlation. J Appl Physiol, 2(1):1–15, 1949.

[11] V. L. Katch. Use of the oxygen-body weight ratio in correlational analyses: spurious correlations and statistical considerations. Med Sci Sports, 5(4):253–7, 1973.

[12] S. Doll, M. Dressen, P. E. Geyer, D. N. Itzhak, C. Braun, S. A. Doppler, F. Meier, M. A. Deutsch, H. Lahm, R. Lange, M. Krane, and M. Mann. Region and cell-type resolved quantitative proteomic map of the human heart. Nat Commun, 8(1):1469, 2017.

[13] G. Banfi, A. Colombini, G. Lombardi, and A. Lubkowska. Metabolic markers in sports medicine. Advances in Clinical Chemistry, Vol 56, 56:1–54, 2012.

[14] J. H. O’Keefe, H. R. Patil, C. J. Lavie, A. Magalski, R. A. Vogel, and P. A. McCullough. Potential adverse cardiovascular effects from excessive endurance exercise. Mayo Clinic Proceedings, 87(6):587–595, 2012.

[15] J. R. Ruiz, M. Joyner, and A. Lucia. Crosstalk opposing view: Prolonged intense exercise does not lead to cardiac damage. J Physiol, 591(Pt 20):4943–5, 2013.

[16] S. Duttaroy, D. Thorell, L. Karlsson, and M. Borjesson. A single-bout of one-hour spinning exercise increases troponin t in healthy subjects. Scand Cardiovasc J, 46(1):2–6, 2012.

[17] Teun van Erp, Carl Foster, and Jos J de Koning. Relationship between various training-load measures in elite cyclists during training, road races, and time trials. International journal of sports physiology and performance, 14:493–500, April 2019.

[18] G. M. Stewart, A. Yamada, L. J. Haseler, J. J. Kavanagh, J. Chan, G. Koerbin, C. Wood, and S. Sabapathy. Influence of exercise intensity and duration on functional and biochemical perturbations in the human heart. J Physiol, 594(11):3031–44, 2016.

[19] D. Mayorga-Vega, P. Aguilar-Soto, and J. Viciana. Criterion-related validity of the 20-m shuttle run test for estimating cardiorespiratory fitness: A meta-analysis. J Sports Sci Med, 14(3):536–47, 2015.

[20] J. Friden, M. Sjostrom, and B. Ekblom. Myofibrillar damage following intense eccentric exercise in man. Int J Sports Med, 4(3):170–6, 1983.

[21] Maximilian Sanno, Steffen Willwacher, Gaspar Epro, and Gert-Peter Brüggemann. Positive work contribution shifts from distal to proximal joints during a prolonged run. Medicine and science in sports and exercise, 50:2507–2517, December 2018.

[22] P. U. Saunders, D. B. Pyne, R. D. Telford, and J. A. Hawley. Factors affecting running economy in trained distance runners. Sports Med, 34(7):465–85, 2004.

[23] G. D. Heise and P. E. Martin. Are variations in running economy in humans associated with ground reaction force characteristics? Eur J Appl Physiol, 84(5):438–42, 2001.

[24] A. Pouliot-Laforte, L. N. Veilleux, F. Rauch, and M. Lemay. Validity of an accelerometer as a vertical ground reaction force measuring device in healthy children and adolescents and in children and adolescents with osteogenesis imperfecta type i. Journal of Musculoskeletal & Neuronal Interactions, 14(2):155–161, 2014.

[25] R. A. Kronmal. Spurious correlation and the fallacy of the ratio standard revisited. Journal of the Royal Statistical Society Series a-Statistics in Society, 156:379–392, 1993.

[26] J. W. McEvoy, Y. Chen, C. E. Ndumele, S. D. Solomon, V. Nambi, C. M. Ballantyne, R. S. Blumenthal, J. Coresh, and E. Selvin. Six-year change in high-sensitivity cardiac troponin t and risk of subsequent coronary heart disease, heart failure, and death. JAMA Cardiol, 1(5):519–28, 2016.

[27] S. Everman, J. W. Farris, R. C. Bay, and J. T. Daniels. Elite distance runners: A 45-year follow-up. Med Sci Sports Exerc, 50(1):73–78, 2018.

